# The cost of inaction to strengthen the resilience of primary health care in Latin America and the Caribbean: a modelling study

**DOI:** 10.1101/2025.03.03.25323291

**Authors:** Tharindu Wickramaarachchi, Nick Scott, Pablo Villalobos Dintrans, Marina Gonzalez-Samano, Manuela Villar Uribe

## Abstract

**Background:** The Latin America and the Caribbean (LAC) region will face future public health emergencies due to pandemics, natural disasters, migration, economic crisis or other unforeseen events. These events disrupt healthcare service coverage with consequences for morbidity, mortality and economic productivity. This study aimed to estimate the health and economic cost of potential future health system shocks, as a proxy for the cost of inaction to strengthen the resilience of primary health care.

**Methods:** For 33 countries in LAC, primary health care shock scenarios were modelled as short-term reductions to the coverage of antenatal care and child health interventions using the Lives Saved Tool, and to family planning services and non-communicable disease management using custom models. Primary health care shocks starting in 2026 and leading to 25-50% coverage reductions (50% being a COVID-19-like disruption) with recovery periods of one to five years were compared to a strengthened primary health care scenario with intervention coverage maintained. Excess deaths and unintended pregnancies were estimated for 2026-2030 and converted to lifetime societal economic costs with 3% per annum discounting based on years of life lost (deaths) and reduced workforce productivity (unintended pregnancies).

**Findings:** Depending on the magnitude and recovery time, the modelled primary health care shocks resulted in an additional 600-3,100 stillbirths, 300-1,400 neonatal deaths, 2,000-10,000 child deaths, 2,200-11,300 maternal deaths, 26,000-131,000 non-communicable disease deaths, and 2.7-14.1 million unintended pregnancies over 2026-2030. This translated to US$7-35 billion in societal economic costs per primary health care shock.

**Interpretation:** Substantive investment in primary health care resilience would be warranted to limit the potential impact of health system shocks on service coverage.

**Funding:** The World Bank.

## Introduction

More than five years since the COVID-19 pandemic declaration, the evidence is clear: COVID-19 had a significant impact on Primary Healthcare Systems (PHC) worldwide.^1^ In addition to the millions of deaths directly caused by COVID-19 globally between 2020-2021, the COVID-19 pandemic led to substantive disruptions to the coverage and quality of healthcare services. For example, there was an observed increase in maternal deaths (odds ratio [OR]=1.37), stillbirths (OR=1.28) and neonatal deaths (OR=1.01) globally during versus before the pandemic^2, 3^ due to reductions in antenatal and delivery care services and reduced healthcare seeking behaviours among pregnant women. Disruptions to healthcare services were most prominent in low- and middle-income countries (LMICs)^4^, related to weaknesses in the health systems^5^.

Latin American and Caribbean (LAC) countries were significantly affected by COVID-19, with excess mortality rates well above the Organisation for Economic Co-operation and Development (OECD) average.^6^ The pandemic exposed weaknesses in health systems and added further strain to countries already facing significant structural challenges in their healthcare systems. Across 14 countries in LAC, an average of 20.4% of households reported experiencing disruptions in healthcare in 2020 (with the largest disruptions reported in Ecuador at 44.9%).^7^ In Peru, 30-40% of health care workers stopped working due to fears for their own health.^7^ In Brazil, all medical procedures underwent significant reductions, including a 42.6% reduction in screening, a 28.9% reduction in diagnostic procedures, and a 42.5% reduction in appointments with physicians.^8^ There were also reductions in health care services targeting children: in 2020, the number of consultations for children reduced by 59-63% in Mexico and 46-69% in Argentina compared to the 2018-19 average,^7^ and in 2021, only 10 of 33 LAC countries met the minimum recommended immunisation levels for DTP (90% target), although 16 countries had achieved this target during 2017-19.^7^ Care management programs for NCDs were also impacted, with a 39% reduction in diabetes blood tests in Argentina and a 41% reduction in Chile in 2020, and a 63% reduction in breast cancer screenings in Brazil compared to the 2018-19 average.

The majority of the burdens associated with health emergencies fall within the remit of PHC.^9, 10^ A robust PHC system, which is people- and community-oriented and offers comprehensive, continuous, and coordinated care, can significantly enhance preparedness and resilience during and after health emergencies.^6^ Regional resolutions by Pan American Health Organization (PAHO) Member States emphasize a PHC approach to address population needs, accelerate pandemic recovery, and move towards universal health care.^11^ The World Bank (WB) has also called for investments in PHC-based systems to prepare for future public health emergencies, protect lives, and foster human capital. The *WB-PAHO Lancet Regional Health Americas Commission on PHC and Resilience* seeks to guide future development of PHC and resilience in the region. Despite efforts to date, there remains a critical need to transform PHC systems in LAC to effectively prevent, respond to and recover from health system shocks, particularly as the region faces risks compounded by weak health systems, fragmentation, constrained budgets, and long-standing inequalities.^11^

Quantifying the health and economic impacts of potential shocks to PHC services can provide rationale for investing in PHC resilience. Previous work has modelled the impact that shocks to specific health care services could have on health outcomes. For example, one study estimated that a 9·8–18·5% reduction in high-impact maternal and child health interventions and a 10% increase in wasting for a six month period could result in 253,500 additional child deaths and 12,200 additional maternal deaths across 118 LMICs.^12^ Another study estimated that reductions to family planning services of different length (3-12 months) and magnitude (5-40%) in 114 LMICs could lead to 325,000 to 15 million additional unintended pregnancies.^13^ In general, while some modelling studies are available that estimate the impact of shocks to health services, they have estimated health impacts only or have estimated health and economic impacts but for a limited subset of interventions. To our knowledge, studies are not available quantifying the health impact of shocks to PHC service coverage, or the economic outcomes associated with these shocks.

The objective of this modelling study was to estimate the health and economic cost of reductions in PHC service coverage due to potential future health system shocks for 33 countries in the LAC region. This evidence can be used as a proxy for the cost of inaction to strengthen PHC, and to provide a rationale to support investment in improving PHC resilience.

## Methods

### Overview

The potential impact of health system shocks (e.g., epidemiological events such as COVID-19, dengue, and Zika; demographic such as migration; natural disasters, including floods, hurricanes, fires, and earthquakes; and economic, such as recessions and economic crisis) on PHC were approximated as reduced service coverage, with the magnitude and duration of the coverage reduction dependent on the nature of the health system shock. The impact of coverage reductions were estimated for family planning, antenatal care, child health, and non-communicable disease (NCD) services domains (Figure 1), compared to a scenario with maintained coverage (“strengthened primary health care”).

**Figure 1:**
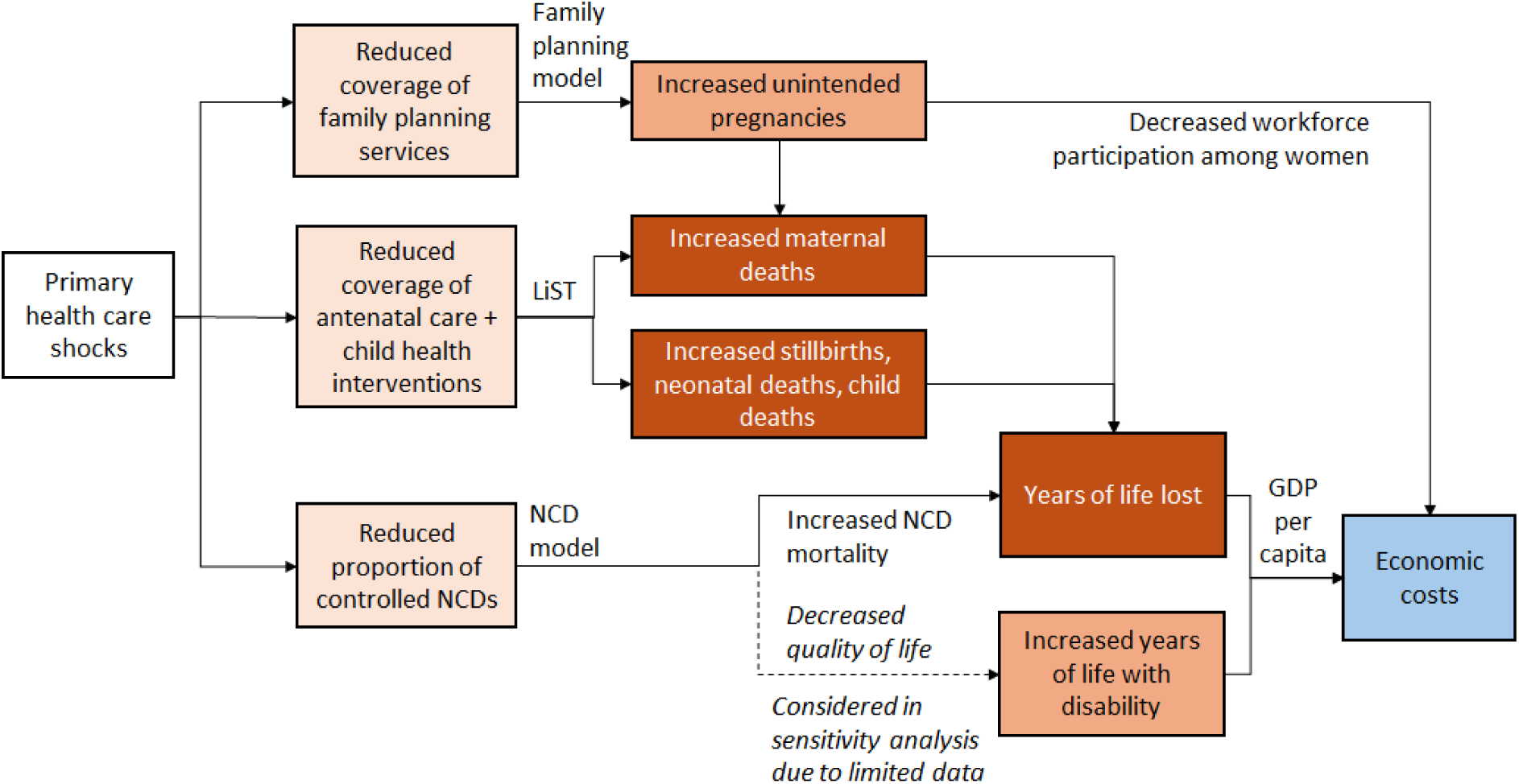
Framework used to estimate the health and economic costs of shocks to primary health care services. The Lives Saved Tool (LiST)^14^ was used to estimate the impact of reduced coverage of antenatal care and child health interventions and custom models were used to estimate the impact of reduced primary health care coverage on family planning and non-communicable disease (NCD) mortality and years lived with disability.

**Figure 2:**
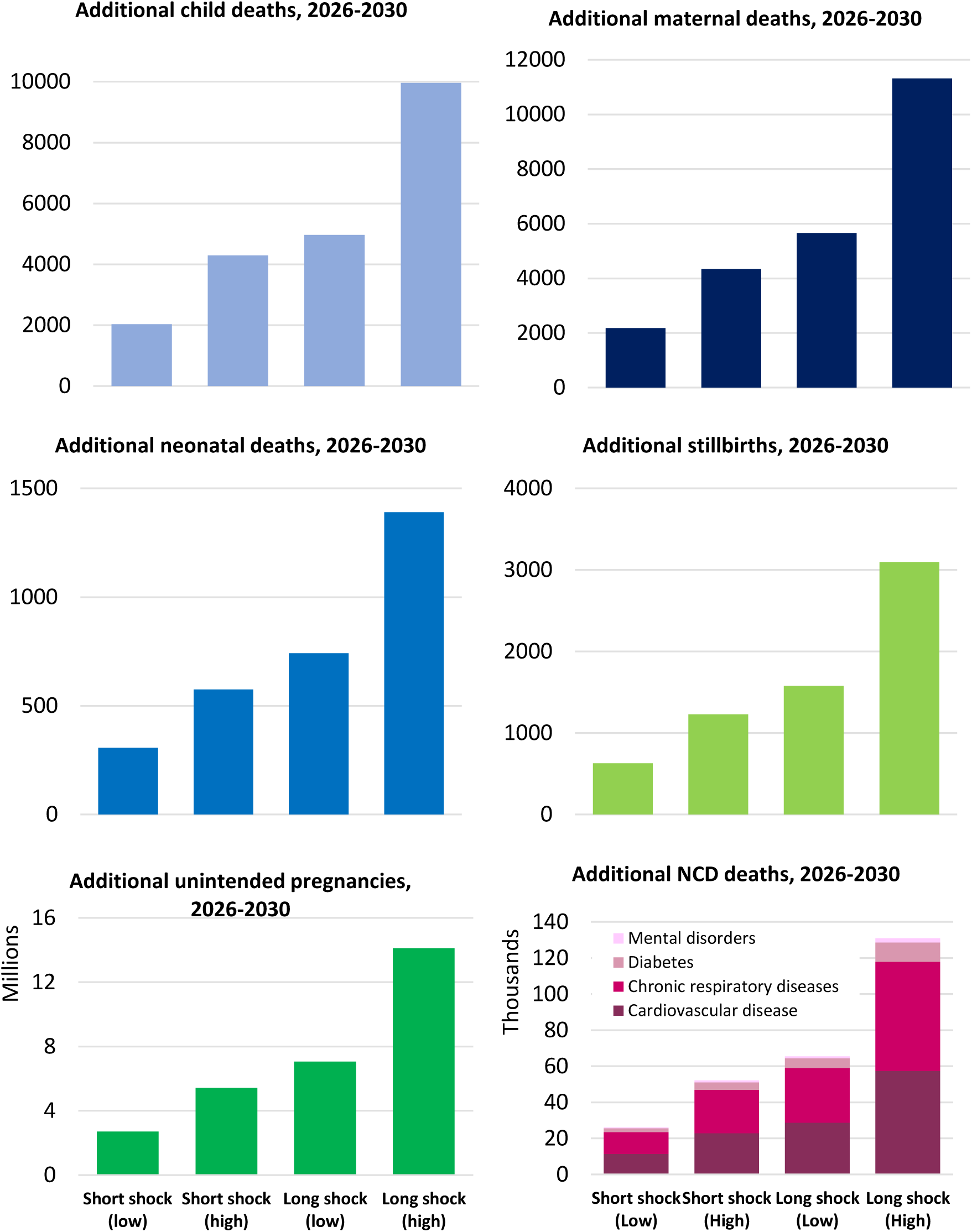
Cumulative additional health outcomes for each primary health care shock scenario. Additional stillbirths (top; left), neonatal deaths (top, middle), NCD deaths (top; right), child deaths (bottom; left), maternal deaths (bottom; middle) and additional unintended pregnancies (bottom; right) over 2026-2030.

### Settings

The modelling was undertaken for 33 countries in the LAC region. Some countries did not have all service domains modelled due to limited data to parametrize the models (Appendix A).

### Model structure

The impact of health system shocks on antenatal care and child health service domains was modelled using the Lives Saved Tool (LiST)^14, 15^, a modelling tool widely used to estimate the health impacts of changes in intervention coverage. A full list of interventions considered for each service domain is available in Appendix B (Table B.1). Reduced antenatal care service utilization and reduced coverage of child health interventions (including interruptions to routine BCG, pentavalent, and polio immunization programs) were modelled to increase maternal deaths, stillbirths, newborn deaths and child deaths relative to the effect sizes in Table 1.

**Table 1:**
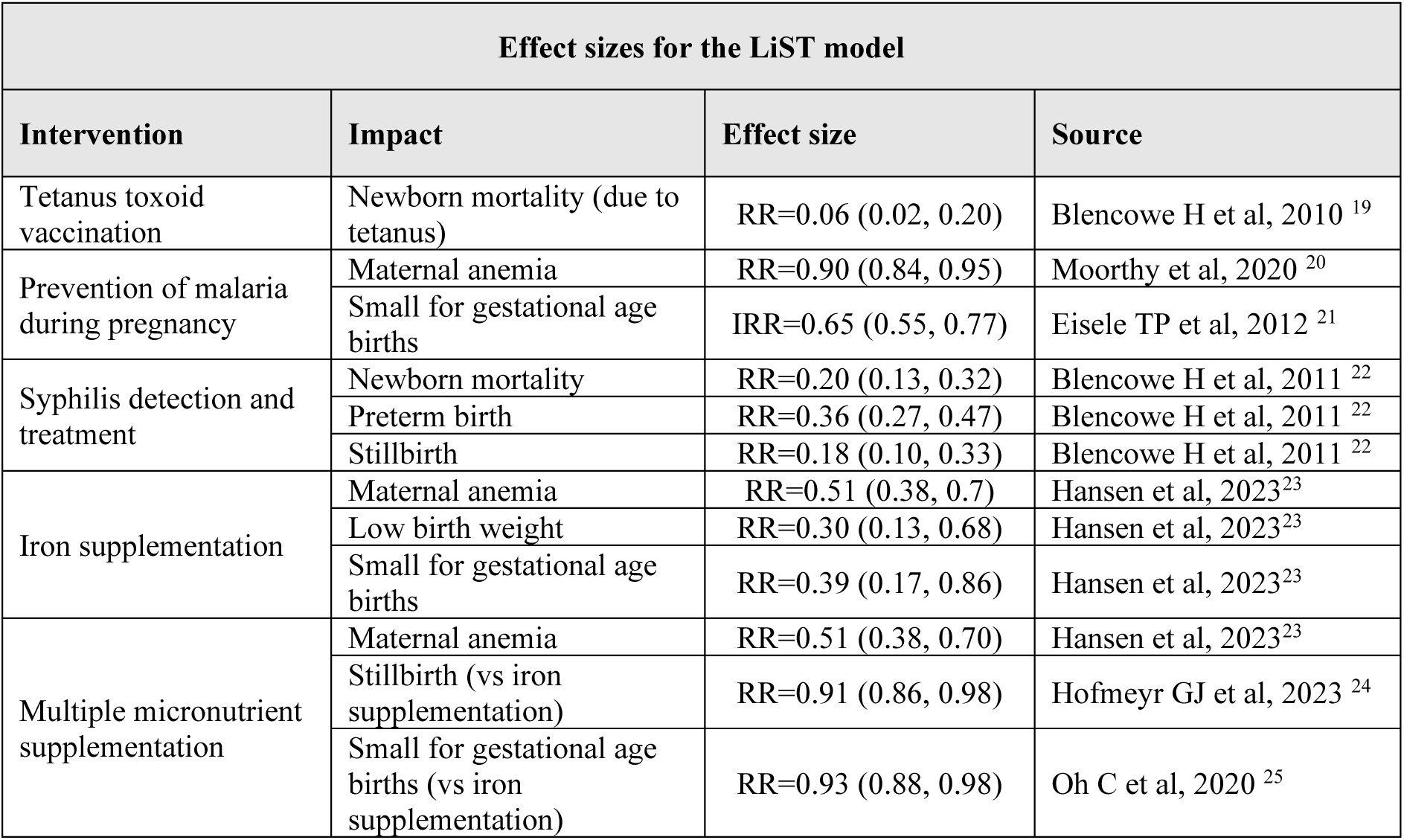
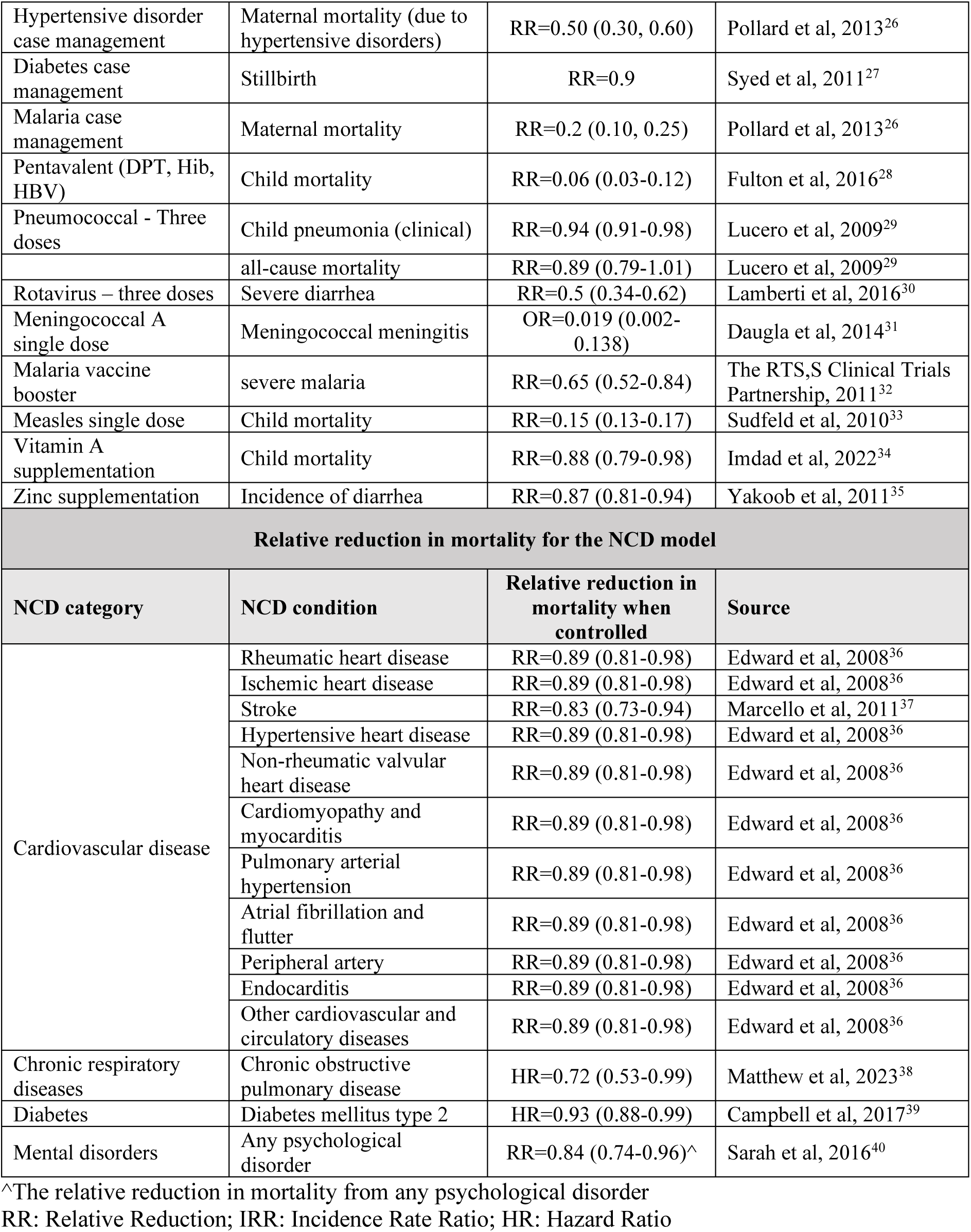
Effect sizes for the interventions modelled in LiST and relative reduction in mortality risk of non-communicable diseases (NCDs)

| Effect sizes for the LiST model |  |  |  |
| --- | --- | --- | --- |
| Intervention | Impact | Effect size | Source |
| Tetanus toxoid vaccination | Newborn mortality (due to tetanus) | RR=0.06 (0.02, 0.20) | Blencowe H et al, 2010 <sup>19</sup> |
| Prevention of malaria during pregnancy | Maternal anemia | RR=0.90 (0.84, 0.95) | Moorthy et al, 2020 <sup>20</sup> |
|  | Small for gestational age births | IRR=0.65 (0.55, 0.77) | Eisele TP et al, 2012 <sup>21</sup> |
| Syphilis detection and treatment | Newborn mortality | RR=0.20 (0.13, 0.32) | Blencowe H et al, 2011 <sup>22</sup> |
|  | Preterm birth | RR=0.36 (0.27, 0.47) | Blencowe H et al, 2011 <sup>22</sup> |
|  | Stillbirth | RR=0.18 (0.10, 0.33) | Blencowe H et al, 2011 <sup>22</sup> |
| Iron supplementation | Maternal anemia | RR=0.51 (0.38, 0.7) | Hansen et al, 2023<sup>23</sup> |
|  | Low birth weight | RR=0.30 (0.13, 0.68) | Hansen et al, 2023<sup>23</sup> |
|  | Small for gestational age births | RR=0.39 (0.17, 0.86) | Hansen et al, 2023<sup>23</sup> |
| Multiple micronutrient supplementation | Maternal anemia | RR=0.51 (0.38, 0.70) | Hansen et al, 2023<sup>23</sup> |
|  | Stillbirth (vs iron supplementation) | RR=0.91 (0.86, 0.98) | Hofmeyr GJ et al, 2023 <sup>24</sup> |
|  | Small for gestational age births (vs iron supplementation) | RR=0.93 (0.88, 0.98) | Oh C et al, 2020 <sup>25</sup> |

| Hypertensive disorder case management | Maternal mortality (due to hypertensive disorders) | RR=0.50 (0.30, 0.60) | Pollard et al, 2013<sup>26</sup> |
| --- | --- | --- | --- |
| Diabetes case management | Stillbirth | RR=0.9 | Syed et al, 2011<sup>27</sup> |
| Malaria case management | Maternal mortality | RR=0.2 (0.10, 0.25) | Pollard et al, 2013<sup>26</sup> |
| Pentavalent (DPT, Hib, HBV) | Child mortality | RR=0.06 (0.03-0.12) | Fulton et al, 2016<sup>28</sup> |
| Pneumococcal - Three doses | Child pneumonia (clinical) | RR=0.94 (0.91-0.98) | Lucero et al, 2009<sup>29</sup> |
|  | all-cause mortality | RR=0.89 (0.79-1.01) | Lucero et al, 2009<sup>29</sup> |
| Rotavirus – three doses | Severe diarrhea | RR=0.5 (0.34-0.62) | Lamberti et al, 2016<sup>30</sup> |
| Meningococcal A single dose | Meningococcal meningitis | OR=0.019 (0.002-0.138) | Daugla et al, 2014<sup>31</sup> |
| Malaria vaccine booster | severe malaria | RR=0.65 (0.52-0.84) | The RTS,S Clinical Trials Partnership, 2011<sup>32</sup> |
| Measles single dose | Child mortality | RR=0.15 (0.13-0.17) | Sudfeld et al, 2010<sup>33</sup> |
| Vitamin A supplementation | Child mortality | RR=0.88 (0.79-0.98) | Imdad et al, 2022<sup>34</sup> |
| Zinc supplementation | Incidence of diarrhea | RR=0.87 (0.81-0.94) | Yakoob et al, 2011<sup>35</sup> |
| **Relative reduction in mortality for the NCD model** |  |  |  |
| **NCD category** | **NCD condition** | **Relative reduction in mortality when controlled** | **Source** |
| Cardiovascular disease | Rheumatic heart disease | RR=0.89 (0.81-0.98) | Edward et al, 2008<sup>36</sup> |
|  | Ischemic heart disease | RR=0.89 (0.81-0.98) | Edward et al, 2008<sup>36</sup> |
|  | Stroke | RR=0.83 (0.73-0.94) | Marcello et al, 2011<sup>37</sup> |
|  | Hypertensive heart disease | RR=0.89 (0.81-0.98) | Edward et al, 2008<sup>36</sup> |
|  | Non-rheumatic valvular heart disease | RR=0.89 (0.81-0.98) | Edward et al, 2008<sup>36</sup> |
|  | Cardiomyopathy and myocarditis | RR=0.89 (0.81-0.98) | Edward et al, 2008<sup>36</sup> |
|  | Pulmonary arterial hypertension | RR=0.89 (0.81-0.98) | Edward et al, 2008<sup>36</sup> |
|  | Atrial fibrillation and flutter | RR=0.89 (0.81-0.98) | Edward et al, 2008<sup>36</sup> |
|  | Peripheral artery | RR=0.89 (0.81-0.98) | Edward et al, 2008<sup>36</sup> |
|  | Endocarditis | RR=0.89 (0.81-0.98) | Edward et al, 2008<sup>36</sup> |
|  | Other cardiovascular and circulatory diseases | RR=0.89 (0.81-0.98) | Edward et al, 2008<sup>36</sup> |
| Chronic respiratory diseases | Chronic obstructive pulmonary disease | HR=0.72 (0.53-0.99) | Matthew et al, 2023<sup>38</sup> |
| Diabetes | Diabetes mellitus type 2 | HR=0.93 (0.88-0.99) | Campbell et al, 2017<sup>39</sup> |
| Mental disorders | Any psychological disorder | RR=0.84 (0.74-0.96)<sup>^</sup> | Sarah et al, 2016<sup>40</sup> |

<sup>^</sup>The relative reduction in mortality from any psychological disorder RR: Relative Reduction; IRR: Incidence Rate Ratio; HR: Hazard Ratio

The impact of health system shocks on family planning services was modelled as a reduction in the proportion of women accessing modern contraception services free of charge (57.8% as a proxy for public vs private access).^16^ Those impacted were modelled to shift from modern methods (average effectiveness 91-99% based on country-specific methods mix; Appendix Table D.3) to traditional methods (average effectiveness of 82%).^14^ This change in contraception effectiveness was modelled to increase unintended pregnancies (accounting for country-specific fertility rates and overall contraception prevalence rates). Additional maternal deaths were also included from the additional pregnancies based on country-specific maternal mortality ratios.

The impact of health system shocks on NCDs was estimated using a custom model, with NCDs across four categories (cardiovascular disease, chronic respiratory disease, diabetes, and mental disorders), selected because they are amenable to PHC as well as the availability of estimates for the relative reduction in mortality when NCDs are controlled (Table 1). Cancers were not included because although reduced screening is likely to lead to missed diagnoses and opportunities for early treatment, it is not clear to what extent a short disruption to screening would be “caught up” following the shock, and whether the role of PHC, as opposed to tertiary care, would be critical in the treatment to prevent cancer-specific mortality. This makes this analysis an underestimate of the impact of health system shocks on NCDs. For each country, the model projected the prevalence of each NCDs and associated age- and cause-specific mortality (based on IHME estimates)^17^, as well as the proportion of people with each NCD accessing primary health care (based on universal health care coverage index)^18^. In the health system shock scenarios, the proportion of people accessing PHC was reduced, leading to increased mortality based on estimates of the impact of primary health care (Table 1).

### Data inputs

The data domains used to populate the models considered for this study are described in Table 2 including their sources. Country-specific values are available in the supplementary material for population projections (Appendix Table D.1 and Table D.2), epidemiological indicators (Appendix Table D.3), NCD prevalence (Appendix Table D.4), NCD mortality rates (Appendix Table D.5), estimated proportion of NCDs that are controlled (Appendix Table D.6), age distribution of NCD deaths (Appendix Table D.7), healthy life expectancy by age (discounted and undiscounted; Appendix Table D.8 and Table D.9), NCD disutility weights (Appendix Table D.10), economic parameters (Appendix Table D.11), and baseline intervention coverages (Appendix Table D.12).

**Table 2:** Main data inputs and their sources.

| Model | Data input | Source |
| --- | --- | --- |
| **LiST model** | Population projections | UN WPP 2024 projections |
|  | Mortality rates: maternal mortality, neonatal mortality, child mortality, stillbirths | WHO Global Health Observatory; WHO Maternal, Newborn, Child and Adolescent Health and Ageing |
|  | Prevalence of stunting and wasting |  |
|  | Contraception prevalence rate |  |
|  | Unmet need for family planning |  |
|  | Baseline coverage of interventions (ANC, vaccines and child health) | LiST v6.36 based on country surveys; regional estimates* |
| **NCD model** | Population projections | UN WPP 2024 projections |
|  | Prevalence of NCD conditions | IHME/GBD<sup>17</sup> |
|  | Mortality rates for NCD conditions | IHME/GBD<sup>17</sup> |
|  | Percentage controlled/managed NCD conditions<sup>^</sup> | Estimated using UHC index and country data<sup>18</sup> |
| **Economic model** | Age- and country-specific healthy life expectancy | IHME/GBD<sup>17</sup> |
|  | Age-distribution of NCD mortality (by cause) | Derived from deaths; IHME/GBD<sup>17</sup> |
|  | GDP per capita | World Bank data<sup>41</sup> |
|  | Female workforce participation rates | World Bank data<sup>42</sup> |
|  | Maternal time out of workforce | UNICEF;<sup>43</sup> The Caribbean Society for Human Resource Professionals<sup>44</sup> |

\*Extracted from LiST; based on country surveys or regional estimates if no survey conducted for the country (Appendix Table D.12)

UHC: Universal Health Coverage; IHME: Institute for Health Metrics and Evaluation; GBD: Global Burden of Disease

**Table 3:**
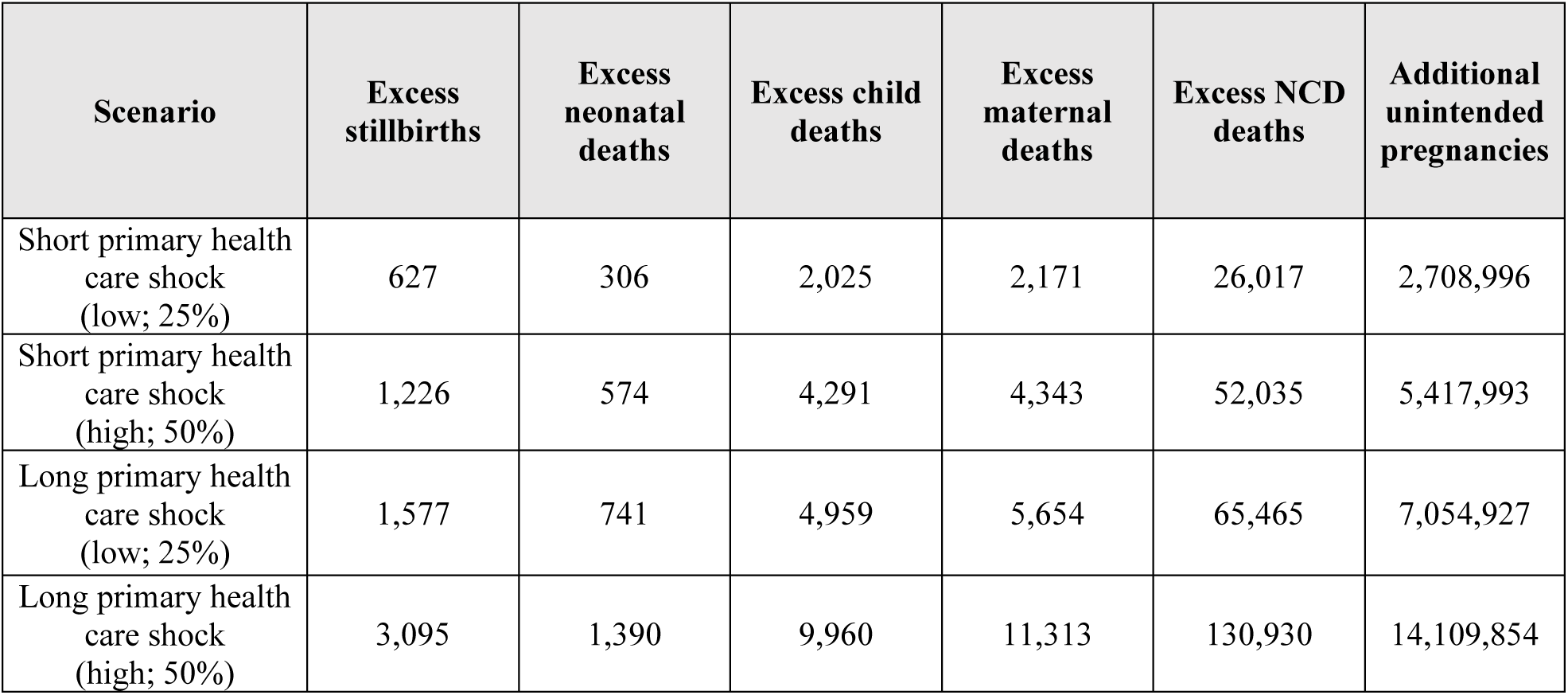
Cumulative health outcomes of primary health care shock scenarios, 2026-2030.

### Scenarios

Assuming that a potential health system shock occurs in 2026, several scenarios were projected for the period 2026-2030:

1. **Strengthened PHC:** Assuming investment occurs to strengthen PHC, such that there are no changes to PHC intervention coverages when the health system shock occurs.
2. **Short PHC shock (low/high)**: PHC intervention coverage in 2026 assumed to reduce by (a) 25% or (b) 50%, before returning to baseline levels in 2027 and maintained through to 2030.
3. **Long PHC shock (low/high)**: PHC intervention coverage in 2026 assumed to reduce by (a) 25% or (b) 50%, before linearly increasing to reach baseline levels again by 2030.

These scenarios were derived such that the PHC shock corresponds approximately to the first year of COVID-19 (for example, in Brazil an 42.5% reduction in appointments for physicians was observed).^8^

### Health impact of PHC shock scenarios

For each scenario, the main outcomes compared to the strengthened PHC scenario were stillbirths, neonatal deaths, child deaths, maternal deaths, deaths due to NCDs and unintended pregnancies over the period of 2026-2030.

### Societal economic costs of PHC shock scenarios

The health impacts of each scenario compared to the strengthened PHC scenario were converted to societal economic costs, using a lifetime time horizon for those impacted across the 2026-2030 period. For each country, excess deaths (maternal, newborn and child deaths, stillbirths, and NCD-related deaths) were converted to years of life lost based on country-specific distributions for the age of different NCD deaths and age of pregnancies, combined with country- and age-specific expected years of life remaining.^45, 46^ Years of life lost were valued at 2023 GDP per capita, with future years discounted at 3% per annum.

Additional unintended pregnancies were assumed to lead to reduced workforce participation, with country-specific maternity leave policies used to estimate time out of the workforce and adjustments made for workforce participation among women.^45, 47^ Time out of the workforce was valued at country-specific GDP per worker, with future years discounted at 3% per annum.

### Sensitivity Analysis

Two sensitivity analyses were run. First, where additional stillbirths and newborn deaths resulting from the additional pregnancies were also considered; these were not included in the main analysis to avoid circular logic in counting the societal economic costs (i.e., in the strengthened scenario their conception did not occur, whereas in the health system shock scenario they contributed many years of life lost), however they would still be expected outcomes. Second, there are likely to be additional impacts from disrupted NCD services leading to increased years lived with disability; however, studies are not available quantifying how NCD disability weights are impacted by accessing PHC. Therefore, a sensitivity analysis was conducted considering the additional economic costs of the health system shock scenarios if accessing PHC led to a 5% reduction in disutility weights for each NCD.

## Results

### Strengthened PHC scenario

In the strengthened PHC scenario with service coverage maintained over 2026-2030, across the 33 countries and the subset of service domains considered, we estimated 367,000 stillbirths, 429,000 neonatal deaths, 327,000 child deaths, 41,000 maternal deaths, ∼7 million deaths from NCDs (deaths due to cardiovascular diseases: 4.5 million; chronic respiratory diseases: 1 million; diabetes: 1.2 million and mental disorders: 211,000) and 57 million births (Appendix Figure F.1).

### PHC shock scenarios: health impacts

Depending on the magnitude (low or high) and duration (short or long) of the shock to PHC services, we estimated an additional 600-3,100 stillbirths, 300-1,400 neonatal deaths, 2,000-10,000 child deaths, 2,100-11,300 maternal deaths, 26,000-131,000 deaths from NCDs and 2.7-14.1 million unintended pregnancies across the 33 countries over 2026-2030 (**Error! Reference source not found.**; **Error! Reference source not found.**).

NCD deaths were the largest share of total excess mortality (83-84%), followed by maternal deaths (7%), child deaths (6-7%), stillbirths (2%) and neonatal deaths (1%).

### PHC shock scenarios: economic impacts

The excess health burden in the shock scenarios corresponded to a total discounted economic cost ranging from US$6.9-35.4 billion (**Error! Reference source not found.**), depending on magnitude and duration of the shock. Reductions in workforce participation due to additional unintended pregnancies represented the largest share of the economic cost (52%), followed by excess deaths due to NCDs (32- 33%) and maternal deaths (7-9%).

Most of the economic costs from excess NCD deaths were from cardiovascular diseases (45%), followed by chronic respiratory diseases (41%), diabetes (10%), and mental disorders (4%). Differences between NCD subcategories were due to prevalence, age-standardized mortality rates and the impact of PHC in reducing mortality (**Table 1**).

### Differences across countries

Out of the total economic cost of the PHC shock scenarios in the LAC, the largest proportion of costs was from Brazil (33%), followed by Mexico (25%), Venezuela (8%), and Argentina (7%), largely driven by population size (**Error! Reference source not found.**). The impact of each shock scenario as a proportion of GDP were more similar across countries (Figure 4), with the short and low magnitude scenario costing between 0.02-0.17% of countries’ GDPs, the short and high magnitude scenario costing between 0.04-0.36% of countries’ GDPs, the long and low magnitude scenario costing between 0.05- 0.44% of countries’ GDPs, and the long and high magnitude scenario costing between 0.11-0.89% of countries’ GDPs. The economic cost of PHC shocks was the highest as a percentage of GDP in Haiti (0.17%-0.89%) followed by Honduras (0.14%-0.72%).

**Figure 3:**
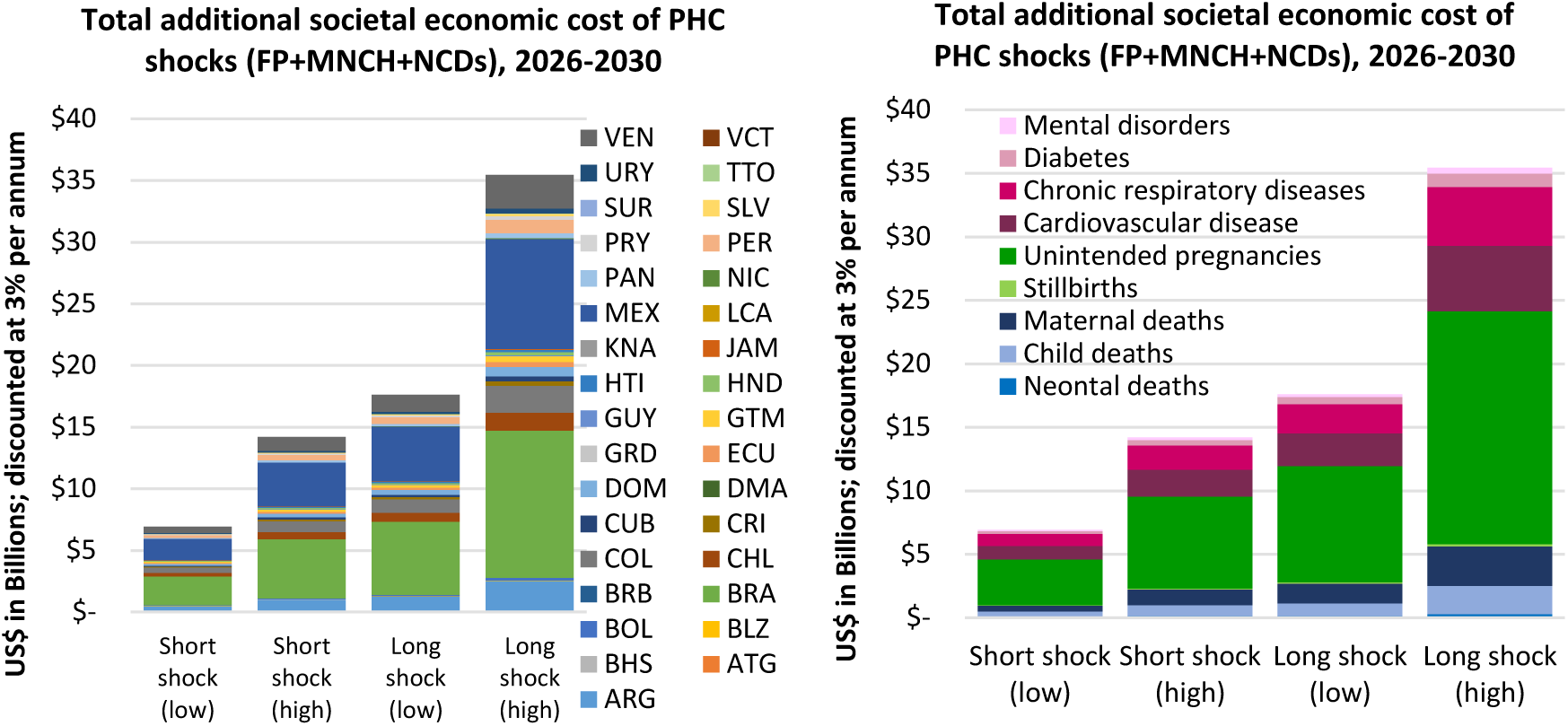
C**u**mulative **societal economic cost of primary health care shock scenarios, disaggregated across countries (left), and across cause of mortality or unintended pregnancies (right).** Costs are total over 2026-2030 across the 33 countries and subsets of service areas modelled, presented in 2023 US$ with 3% per annum discounting applied. The economic costs vary across countries due to relative population size and epidemiological indicators. FP = family planning; MNCH = maternal, newborn and child health; NCDs = non-communicable diseases.

**Figure 4:**
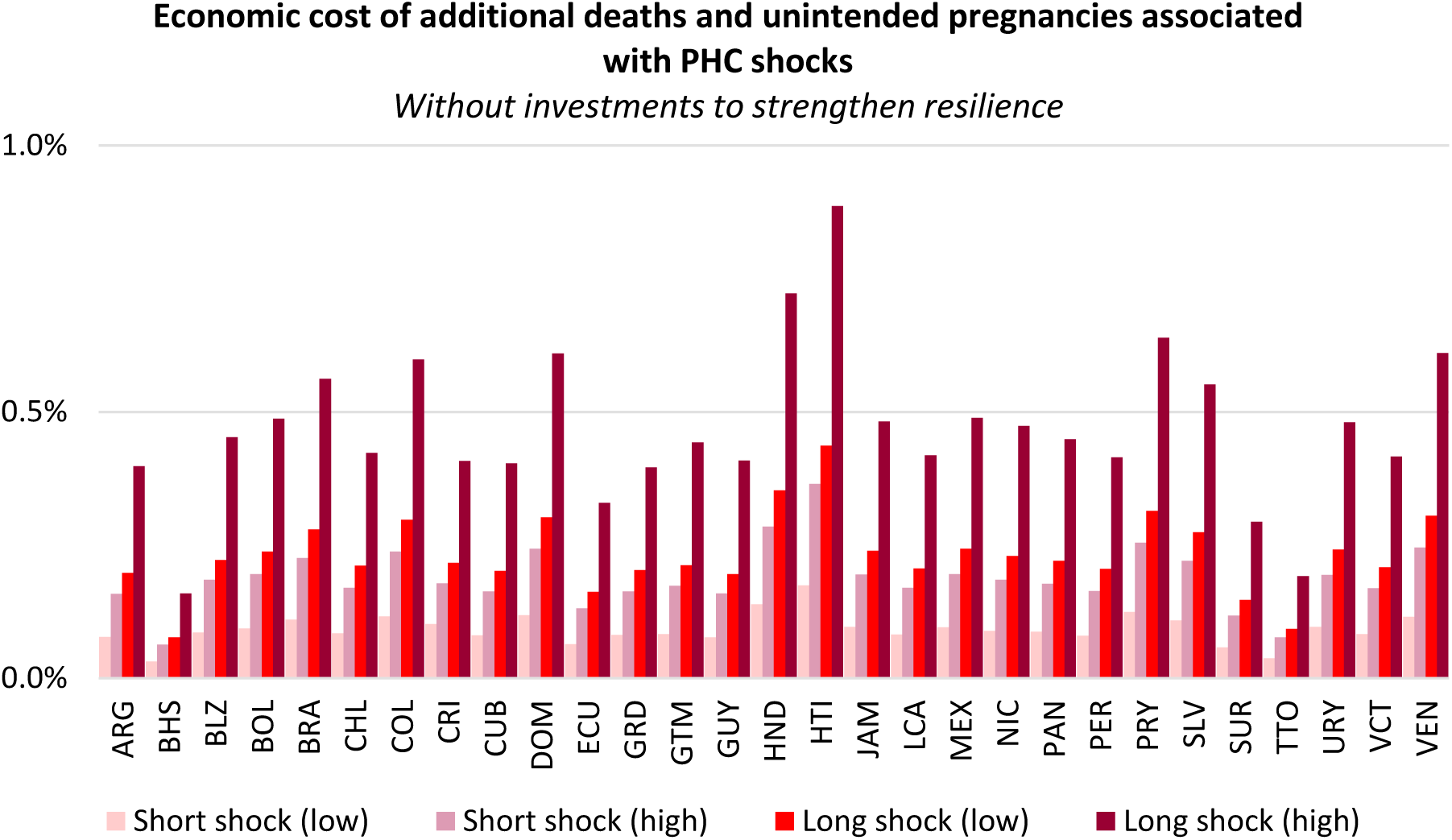
E**c**onomic **cost of primary health care shocks as a percentage of the national GDP.** 2023 GDP values were used; the economic cost in some countries may appear small relative to population size as not all intervention categories could be included for some of the smaller countries (Appendix A). The economic costs of the shocks occur spread over many years but totals are shown as a percentage of annual GDP for contextualisation.

### Sensitivity analyses

When additional stillbirths and neonatal deaths were considered as a result of the additional pregnancies (using country-specific stillbirth and neonatal mortality rates), this increased total stillbirths in the PHC shock scenarios from 600-3,100 to 20,000-105,000 and neonatal deaths from 300-1,400 to 23,000- 122,000 (Appendix Figure F.2). This increased the economic cost of the PHC shock scenarios from US$6.9-35.4 billion to US$14.8-75.1 billion (Appendix Figure F.3). Including an assumed 5% reduction in disutility when NCD conditions are controlled increased the economic cost of the shock scenarios from US$6.9-35.4 billion to US$12.3-61.9 billion over 2026-2030 (Appendix Figure F.4).

## Discussion

This study estimated the health and economic cost of potential future public health emergencies or large-scale shocks to PHC in the LAC region, as a proxy for the cost of inaction to strengthen PHC. The results suggest that events that result in 25-50% reductions in PHC coverage, with recovery time ranging from one to five years, could lead to an additional 31,000-157,000 deaths (stillbirths, neonatal deaths, child deaths, maternal deaths and deaths due to NCDs), and 2.7-14.1 million unintended pregnancies across the 33 countries in the LAC region. These health impacts translate to cumulative societal economic costs of US$6.9-35.4 billion due to years of life lost and reduced workforce participation.

It is unknown when or what type of health system shocks will occur in the future, and so the range of scenarios considered have been designed to reflect a range of potential shocks. To put these in context, the large magnitude PHC shock scenarios are roughly consistent with observations reported in the LAC region during the COVID-19 pandemic. For example, in Brazil, which represents nearly one third of the population in the region, there was a 42.5% reduction in appointments with physicians^8^ and in Mexico and Argentina there were 59-63% and 46-69% reductions in consultations with children, respectively.^7^ Natural disasters or mass population movements could also have large effects; in 2018, the government of Colombia granted nearly 477,000 special stay permits to Venezuelan citizens in response to a mass migration.^48^ Some events could cross territorial borders: during economic and political crises in Venezuela, malaria cases resurged and neighbouring countries, such as Brazil, reported an increasing trend of imported cases .^49^ It is also possible that multiple health system shocks could occur together and in sequence, which would put further strain on the health system if proper planning and strengthening has not occurred.

The estimated societal economic costs of the PHC shock scenarios represent a lower bound, as they are based on a selection of services delivered through PHC only, and many additional impacts were not accounted for. First, the NCD model only captures mortality and not morbidity. This is because while disutility weights are available for different NCDs, there is limited evidence on the relative reduction in these disutility weights when PHC is accessed and/or the conditions are controlled. In a sensitivity analysis, the total economic cost of the PHC shock scenarios increased to US$12.3-61.9 billion when a 5% reduction in NCD disutility weights was assumed with PHC access (Appendix Figure F.4). Second, only a subset of NCDs were considered, based on the availability of parameters to approximate the impact of PHC on mortality risk (Table 1), and in particular this did not include screening for cancers. Third, other communicable diseases such as HIV, hepatitis and tuberculosis can be managed through PHC and so would also be impacted by shocks to PHC but were not included. Fourth, the estimated costs of PHC shocks were intended as a proxy for the cost of inaction, however investment to strengthen PHC could have additional benefits even if shocks do not occur, which are not captured in this study. Fifth, in the main analysis excess stillbirths and neonatal deaths resulting from additional unintended pregnancies were not included; in a sensitivity analysis their inclusion more than doubled the total economic cost of the PHC shock scenarios (from US$6.9-35.4 billion to US$14.8-75.1; Appendix Figure F.3).

Most of the total impact came from countries with the largest population sizes; however, as a proportion of GDP the relative impacts were similar in each country. For example, Brazil, Mexico, Venezuela and Colombia represent 64% of the population from the 33 countries and accounted for 73% of the total economic costs of the PHC shock scenarios. As a proportion of GDP, the societal cost of these shock scenarios ranged from 0.02-0.17% in the short and low magnitude scenario to 0.11-0.89% in the long and high magnitude scenario. It is difficult to benchmark these estimates against data from COVID-19, where for example during the first year of COVID-19 pandemic aggregate GDP contracted by 7% in the LAC region,^7^ since this observed 7% includes the direct impacts of COVID-19, shocks to health services beyond PHC, and shocks to all other sectors.

Some components of this work can be compared against estimates relating service disruptions from COVID-19 to health outcomes. For example, Lara et al.^50^ estimated that a 28% reduction in non-COVID-19-related hospitalisations in Brazil, Ecuador, Mexico, and Peru could lead to a 15% increase in mortality rates compared with pre-pandemic years, with 89% of the mortality increase due to NCDs. When considering our short and low magnitude PHC shock scenario (25% reduction in PHC for one year) in just these four countries, and only considering maternal, newborn and NCD-related deaths to align with Lara et al.^50^, our model estimates an excess ∼18,000 deaths (an ∼0.25% increase compared to no shock). However, it should be noted that Lara et al. consider hospitalisations for which health system shocks are more likely to have an immediate impact of mortality, whereas this study is considering PHC-delivered interventions only. In another study, UNFPA estimated that across 33 countries in the LAC region, a six-month moderate reduction to public sector utilization of modern contraceptives services could have led to 1.7 million additional unintended pregnancies, 2,900 maternal deaths and 39,000 infant deaths.^51^ This is aligned with our modelled estimates of PHC shocks over a one to five year time frame leading to 2.7-14.1 million unintended pregnancies and 2,100-11,300 maternal deaths.

This study did not consider actions required to strengthen PHC, but investment in PHC resilience will be necessary to reduce the impact of external shocks in the future, while also enhancing health services even in the absence of these events. It has been recommended that OECD countries invest 1.4% of GDP annually on average to strengthen health system resilience overall, based on 2019 expenditure, in health workforce, prevention, and key infrastructure.^52^ This includes supporting frontline health professionals (at 0.7% of GDP) and additional investments in preventive care (at 0.3% of GDP), which is broadly consistent with the recent WHO recommendation for countries to allocate extra 1% of GDP targeting PHC.^52^ Policy interventions to expand fiscal space, improve revenue collection, enhance budgeting practices, optimize pooling funds and resource allocations, and establish efficient purchasing and payment systems for providers will be critical.^7, 52^ Programmatically, system fragmentation will need to be reduced, and the benefits packages offered by PHC clearly defined. Infrastructure will also be required, including to strengthen the capacity of supply chains.^6, 52^ This will take coordinated efforts across various government levels and sectors. Finally, accountability, transparency, and equitable resource allocations will be vital to ensure the long-term sustainability of PHC.

There are several limitations to this study. First, demographic and epidemiological data to parametrize the models for each country and service area were taken from a variety of sources, each with their own bias. Where possible, primary sources such as Demographic and Health Services or other large surveys were used, but in many cases model inputs were taken from the LiST model or Institute of Health Metrics and Evaluation databases (Appendix D), which are imputed or are the outputs of other models. In addition, the relative Universal Health Coverage service index^18^ for each country was used as a proxy for both PHC coverage and the percentage of people with each NCD who had their disease “controlled” (Appendix Table D.6); it is unclear whether this is likely to be an underestimate or overestimate. Second, not all countries had all service domains considered in the analysis. For example, there was not enough data to model shocks to family planning, ANC, and vaccination services for 4/33 countries (12%), child health services for 21/33 countries (64%), and other pregnancy-related services for 18/33 countries (55%) (Appendix A); however, the missing data was mostly from countries with small populations and so is unlikely to have major impacts to the aggregate outcomes. Third, the cause, timing and magnitude of future shocks is unknown, and so the PHC shock scenarios are theoretical and meant to reflect a range of possibilities rather than being specific to any type of event. We also modelled the shocks to begin in 2026 because the most recent data for each country is more reflective of 2026 than of future years, however in the absence of major changes to epidemiological indicators and interventions, the shocks are likely to have similar impacts if they occurred in future years. Fourth, the cost of strengthening PHC were not included in this modelling because they are currently unknown but should be a subject of further work.

## Conclusion

The COVID-19 pandemic led to reductions in health service coverage and highlighted weaknesses in PHC systems in the LAC region. If another health system shock occurred that led to a 25-50% reduction in PHC service coverage with recovery time ranging from one to five years, then across the 33 countries in the LAC region this could lead to an additional 31,000-157,000 deaths (stillbirths, neonatal deaths, child deaths, maternal deaths and deaths due to NCDs) and 2.7-14.1 million unintended pregnancies, corresponding to US$6.9-35.4 billion in societal economic costs. Therefore, substantive investment in PHC resilience would be warranted if it could limit or eliminate the impact of shocks on service coverage.

## Supporting information

Supplementary methods and results

Data sets

## Data Availability

All data produced in the present work are contained in the manuscript and in its supplements.

## Acknowledgements

The authors would like to acknowledge the feedback and inputs from Marcia Castro, Sebastian Bauhoff, James Macinko, Frederico Guanais, Sara Bennett, and Cristian Herrera.

## Author contributions

All authors conceived the study. TW and NS designed the model with inputs from PAVD, RMGS, MVU. TW implemented model scenarios. TW and NS analysed model outputs. PAVD, RMGS, MVU validated data inputs. TW and NS drafted the manuscript. All authors contributed to the final version of the manuscript.

