## Supplementary methods and results for "The cost of inaction to strengthen the resilience of primary health care in Latin America and the Caribbean: a modelling study"

### Appendices

#### Appendix A: Data availability for countries across different service domains

**Table A.1: Service coverage data availability across countries**

| ISO | Family planning | ANC visits | Pregnancy related routines and maternal nutrition | Vaccines | Child health | NCD |
|---|---|---|---|---|---|---|
| ARG | ✓ | ✓ | X | ✓ | X | ✓ |
| ATG | X | X | X | X | X | ✓ |
| BHS | ✓ | ✓ | X | ✓ | X | ✓ |
| BLZ | ✓ | ✓ | ✓ | ✓ | ✓ | ✓ |
| BOL | ✓ | ✓ | ✓ | ✓ | ✓ | ✓ |
| BRA | ✓ | ✓ | ✓ | ✓ | X | ✓ |
| BRB | X | X | X | X | X | ✓ |
| CHL | ✓ | ✓ | X | ✓ | X | ✓ |
| COL | ✓ | ✓ | ✓ | ✓ | X | ✓ |
| CRI | ✓ | ✓ | X | ✓ | X | ✓ |
| CUB | ✓ | ✓ | X | ✓ | X | ✓ |
| DMA | X | X | X | X | X | ✓ |
| DOM | ✓ | ✓ | ✓ | ✓ | ✓ | ✓ |
| ECU | ✓ | ✓ | ✓ | ✓ | X | ✓ |
| GRD | ✓ | ✓ | X | ✓ | X | ✓ |
| GTM | ✓ | ✓ | ✓ | ✓ | ✓ | ✓ |
| GUY | ✓ | ✓ | ✓ | ✓ | X | ✓ |
| HND | ✓ | ✓ | ✓ | ✓ | ✓ | ✓ |
| HTI | ✓ | ✓ | ✓ | ✓ | ✓ | ✓ |
| JAM | ✓ | ✓ | X | ✓ | X | ✓ |
| KNA | X | X | X | X | X | ✓ |
| LCA | ✓ | ✓ | X | ✓ | X | ✓ |
| MEX | ✓ | ✓ | ✓ | ✓ | ✓ | ✓ |
| NIC | ✓ | ✓ | ✓ | ✓ | ✓ | ✓ |
| PAN | ✓ | ✓ | X | ✓ | ✓ | ✓ |
| PER | ✓ | ✓ | ✓ | ✓ | ✓ | ✓ |
| PRY | ✓ | ✓ | X | ✓ | ✓ | ✓ |
| SLV | ✓ | ✓ | ✓ | ✓ | ✓ | ✓ |
| SUR | ✓ | ✓ | X | ✓ | X | ✓ |
| TTO | ✓ | ✓ | X | ✓ | X | ✓ |
| URY | ✓ | ✓ | X | ✓ | X | ✓ |
| VCT | ✓ | ✓ | X | ✓ | X | ✓ |
| VEN | ✓ | ✓ | ✓ | ✓ | X | ✓ |

### Appendix B: Interventions and NCDs included in the modelling

**Table B.1: Interventions considered in LiST based model**

| Category | Intervention |
|---|---|
| **Antenatal Care** | ANC (1 visit) |
| | ANC (>4 visits) |
| | Tetanus toxoid vaccination |
| | Prevention of malaria during pregnancy |
| | Syphilis detection and treatment* |
| | Iron supplementation |
| | Multiple micronutrient supplementation |
| | Hypertensive disorder case management* |
| | Diabetes case management* |
| | Malaria case management* |
| **Vaccines** | Pentavalent (DPT, Hib, HBV) |
| | Pneumococcal - Three doses |
| | Rotavirus – three doses |
| | Meningococcal A single dose |
| | Malaria vaccine booster |
| | Measles single dose |
| | Measles two doses |
| **Child health** | Vitamin A supplementation |
| | Zinc supplementation |

*Coverage for these pregnancy interventions are calculated in LiST as proportional to ANC utilization multiplied by an estimate for the quality of ANC (refer appendix C)

**Table B.2: NCD conditions considered for modelling**

| Cardiovascular disease | Diabetes | Chronic respiratory disease | Mental disorders |
|---|---|---|---|
| Rheumatic heart disease | Diabetes mellitus type 2 | Chronic obstructive pulmonary disease | Any psychological disorder |
| Ischemic heart disease | | | |
| Stroke | | | |
| Hypertensive heart disorder | | | |
| Non-rheum valve diseases | | | |
| Cardiomyopathy | | | |
| Pulmonary arterial hypertension | | | |
| Atrial fibrillation | | | |
| Peripheral artery | | | |
| Endocarditis | | | |
| Other cardiovascular | | | |

#### Appendix C: Estimating the coverage based on utilization and quality

Ten antenatal care interventions were included in this study from LiST (Table B.1). Of these, Syphilis detection and treatment, hypertensive disorders case management, diabetes case management and malaria case management do not have data on coverage from household surveys. The coverages of these interventions are estimated using service utilization and quality (Table C.1).

**Table C.1. Antenatal care interventions estimated from utilization and quality**

| Intervention | Default data source for coverage |
|---|---|
| Syphilis detection and treatment | Calculated from utilization (at least 1 ANC visit) and quality |
| Hypertensive disorders case management | Calculated from utilization (at least 4 ANC visit) and quality |
| Diabetes case management | Calculated from utilization (at least 4 ANC visit) and quality |
| Malaria case management | Calculated from utilization (at least 4 ANC visit) and quality |

*B.2.1 Quality/readiness-adjusted coverage*

For the interventions in Table C.1, LiST estimates coverage by multiplying utilization by quality/readiness of clinics to provide that service:

$$\text{Coverage estimates} = \text{Quality} \times \text{Utilization}$$

For antenatal care interventions, utilization is based on the attendance to antenatal care clinics (either at least 1 ANC visit, or at least 4 ANC visits; Table C.1). Major household surveys conducted in countries are used to measure the utilization matrices of antenatal care (e.g. DHS, MICS). Quality is a consideration that impacts overall coverage, modifying for the proportion of women who visit antenatal care. Facility survey programs such as Service Provision Assessments (SPA) and Service Availability and Readiness Assessment (SARA) collect data on the services different health facilities are able to provide.[1] In these surveys, drugs, supplies equipment and tests available at the clinics are recorded in addition to the checks on training and supervision of service providers.[1]

#### Appendix D: Data inputs

See supplementary Excel file.

Table D.1: Population projections for countries in the LAC (Totals)

Table D.2: Population estimations for countries in the LAC (for age bins; by sex)

Table D.3: Epidemiological indicator data

Table D.4: Prevalence rates of NCD conditions

Table D.5: Mortality rates for NCD conditions

Table D.6: Percentage of population controlled their NCD conditions (estimated)

Table D.7: Death distribution of NCDs according to five-year age bins

Table D.8: Expected healthy life years for countries in LAC (discounted)

Table D.9: Expected healthy life years for countries in LAC (undiscounted)

Table D.10: Relative reduction in disutility weight if the NCD condition is controlled

Table D.11: Parameters for the economic evaluations

Table D.12: Percentage of ANC utilization and coverages of interventions (pregnancy, child health and vaccines)

#### Appendix E: Economic model description

The economic cost of shocks to primary health care service coverage (antenatal care, prevention, family planning services and management of non-communicable diseases) were considered across two domains:

1. **Social**: increased mortality (increased maternal deaths, stillbirths, neonatal deaths and child deaths) and increased morbidity (years of life with disability due to uncontrolled non-communicable diseases) leading to economic costs
2. **Workforce participation**: reduced workforce participation due to unintended pregnancies

Economic costs of primary health care shocks were calculated based on the difference in outcomes between the strengthened scenario (assuming no shocks to primary health services delivery) compared with each health system shock scenario specified by its duration and the intensity.

*Social costs from years of life lost*

The Lives Saved Tool (LiST) was used to model reduced coverage of antenatal care interventions due to shocks and calculate the additional number of maternal deaths, stillbirths, neonatal deaths and child deaths for each year. Decreased coverage of family planning was also modelled separately, resulting in an increased number of unintended pregnancies as well as additional maternal deaths, which were calculated according to the maternal mortality rates among current pregnancies for each country.

Additional maternal deaths, stillbirths, neonatal deaths and child deaths were converted to total years of life lost using country- and age-specific distributions of expected healthy life years remaining (Table D.8). Stillbirths and neonatal deaths were assumed to have years of life loss equivalent to the expected number of healthy life years of newborns and child deaths were assumed to have years of life loss equivalent to the expected number of healthy life years of a child less than five years old. Country-specific distributions for the age of pregnancies were used to estimate the age of maternal deaths and corresponding expected number of healthy life years lost. There is some debate about years of life gained from averting stillbirths (more specifically disability-adjusted life years gained)[2], and for this analysis years of life lost were considered for 19.7% of the stillbirths occurred, which is the estimate for the Latin America region that are intrapartum[3].

Primary health care shocks could also result in reduced management of non-communicable diseases (NCD) leading to additional deaths. For each NCD, the country-specific age distribution of deaths was used to estimate expected healthy life years lost.

GDP per capita for each country was used to convert years of life lost into economic costs, with 3% per annum discounting applied to future years.

*Social costs from years lived with disability*

The main analysis of this study did not consider years lived with disability. However, in a sensitivity analyses they were considered: a prevalence-based approach was used to calculate the years lived with disability[4], assuming that people who access primary health care have reduced disutility weights for each NCD. Due to primary health care shocks, a greater proportion of people would move to experiencing the higher disutility weight, leading to an increase in total years lived with disability.

*Costs of reduced workforce participation due to unintended pregnancies*

Additional unintended pregnancies occurring among women >18 years due to primary health care shocks were assumed to lead to a reduction in workforce participation. Pregnancy was assumed to remove a woman from the labour force for several months, based on country-specific maternity leave policies (Table D.11) and adjusted for country specific workforce participation rates among women.

The economic cost was then calculated as the duration of maternity leave multiplied by the GDP per worker.

### Appendix F: Additional results

*Total outcomes*

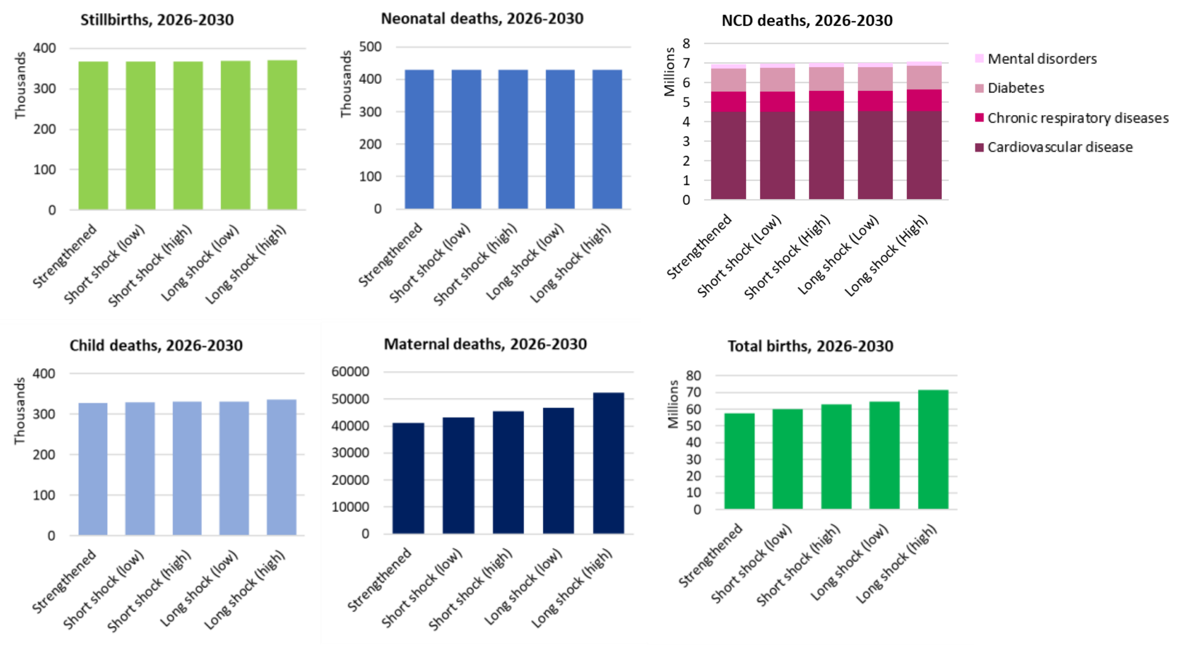


**Figure F.1:** Cumulative total health outcomes for each shock scenario compared with "strengthened" scenario. Total stillbirths (top; left), neonatal deaths (top, middle), NCD deaths (top; right), child deaths (bottom; left), maternal deaths (bottom; middle) and births (bottom; right) over 2026-2030

*Sensitivity analysis: including stillbirths and neonatal deaths due to shocks in family planning services*

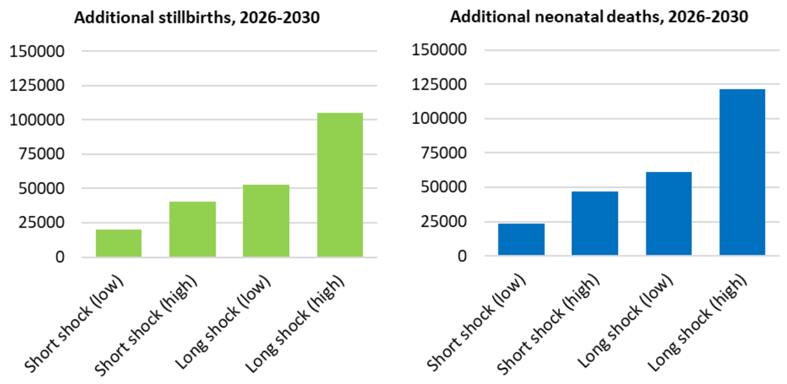


**Figure F.2:** Cumulative stillbirths (left) and neonatal deaths (right) due to shocks in maternal health as well as family planning services

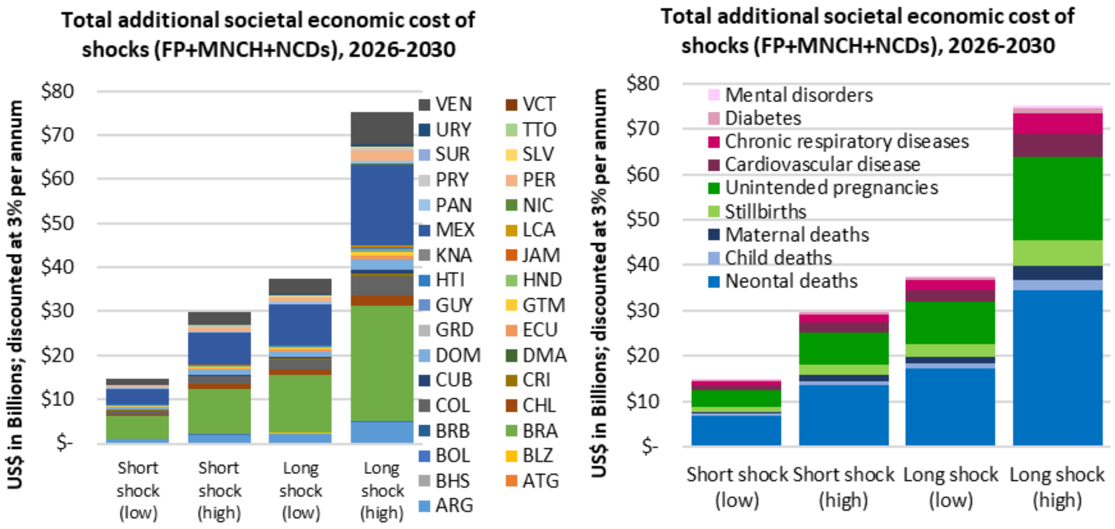


**Figure F.3:** Cumulative societal economic cost of primary health care shock scenarios, disaggregated across countries (left), and across cause of mortality or unintended pregnancies (right). **In this figure, the impact of shocks to family planning services on stillbirths and neonatal deaths is also included.** Costs are total over 2026-2030 across the 33 countries and subsets of service areas modelled, presented in 2023 US$ with 3% per annum discounting applied. The economic costs vary across countries due to relative population size and epidemiological indicators.

*Sensitivity analysis: including a 5% reduction in disutility weight when accessing primary health care*

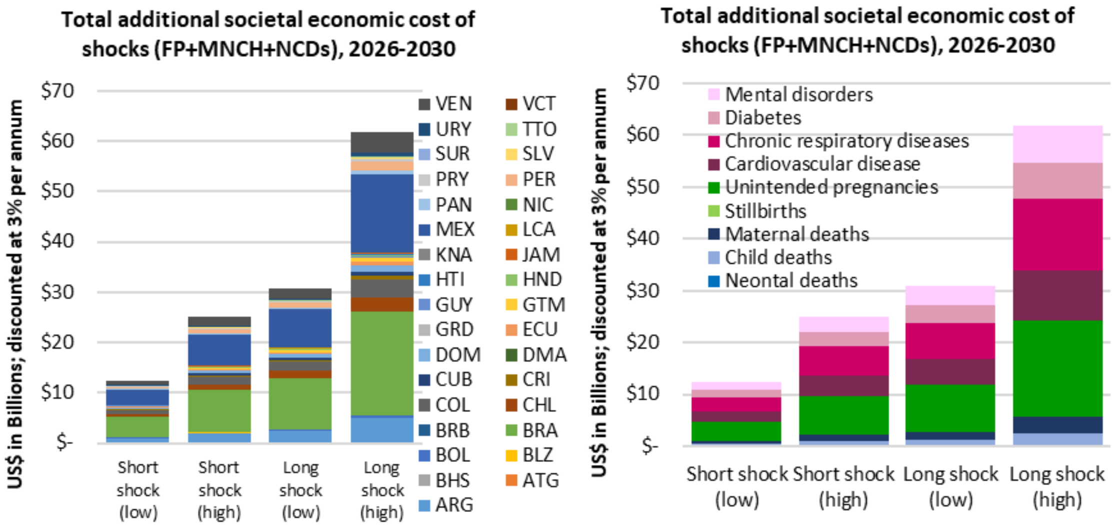


**Figure F.4:** Cumulative societal economic cost of primary health care shock scenarios, disaggregated across countries (left), and across cause of mortality or unintended pregnancies (right). **An assumed 5% reduction in disutility when NCD conditions are controlled included**. Costs are total over 2026-

2030 across the 33 countries and subsets of service areas modelled, presented in 2023 US$ with 3% per annum discounting applied. The economic cost**s** vary across countries due to relative population size and epidemiological indicators.
